# A randomized clinical trial reveals effects of mindfulness and slow breathing on plasma amyloid beta levels

**DOI:** 10.1101/2025.03.10.25323695

**Authors:** Kaoru Nashiro, B. Rael Cahn, Paul Choi, Hye Rynn J. Lee, Shaakhini Satchi, Jungwon Min, Hyun Joo Yoo, Christine Cho, Noah Mercer, Lorena Sordo, Elizabeth Head, Jeiran Choupan, Mara Mather

## Abstract

Prior research suggests that meditation may slow brain aging and reduce the risk of Alzheimer’s disease (AD). However, we lack research systematically examining what aspect(s) of meditation may drive such benefits. In particular, it is unknown how breathing patterns during meditation might influence health outcomes associated with AD. In this study, we examined whether two types of mindfulness meditation practice - one with slow breathing and one with normal breathing – differently affect plasma amyloid beta (Aβ) relative to a no-intervention control group. One week of daily mindfulness practice with slow breathing decreased plasma Aβ levels whereas one week of daily mindfulness practice with normal breathing increased plasma Aβ levels. The no-intervention control group showed no changes in plasma Aβ levels. Slow breathing appears to be a factor through which meditative practices can influence pathways relevant for AD.

**Research Transparency Statement:** Conflicts of interest: All authors declare no conflicts of interest. Funding: This study was supported by Epstein Breakthrough Alzheimer’s Research Fund (PI: Mather, co-PI: Choupan) and by R01AG080652 (PI: Mather). Artificial intelligence: No artificial intelligence assisted technologies were used in this research or the creation of this article. Ethics: This research received approval from the University of Southern California Institutional Review Board (ID: UP-23-00373).

---

Meditative practices have been shown to be associated with slowing brain aging (Lazar et al., 2005; Luders, 2014; Luders et al., 2011, 2014, 2016; Pagnoni & Cekic, 2007) and there is interest in how they may help reduce the risk of or delay the onset of Alzheimer’s disease (AD; (Epel et al., 2016; Innes & Selfe, 2014; Lutz et al., 2021; National Academies of Sciences Engineering and Medicine et al., 2022). While most prior studies focus on how meditative practices benefit health via mental training aspects (e.g., paying attention to the present moment; Chételat et al., 2018; Klimecki et al., 2019; Lutz et al., 2021), with particular emphasis on awareness and acceptance instead of awareness alone(Lindsay et al., 2018; Lindsay & Creswell, 2017), or by promoting relaxation and rest (Crosswell et al., 2024), the effects of different breathing patterns during meditative practices on health have been relatively little explored. Importantly, meditative practices do tend to affect breathing and breath rate (Karunarathne et al., 2023, 2024; Wielgosz et al., 2016), which can elicit powerful physiological effects that might influence AD-related health outcomes.

Separately from meditation per se, slow breathing induces large heart rate oscillations at the breathing frequency (Bernardi et al., 2001; P. Lehrer et al., 1999; C.-K. Peng et al., 2004). Heart rate increases during inhalation and decreases during exhalation. Thus, heart rate oscillates at the breathing frequency, which is usually around 3-4 s per breath (Cysarz & Büssing, 2005; Nashiro et al., 2023; Vai et al., 1988). The amplitude of heart rate oscillation increases as breathing slows, peaking at either around a 10-s (0.1 Hz) pace (Hirsch & Bishop, 1981) or a 15-s (0.67 Hz) pace (H.-S. Song & Lehrer, 2003). Thus, one can dramatically increase one’s own heart rate oscillatory activity by breathing slowly. HRV biofeedback was developed as a method to increase HRV through slow-paced breathing and real-time feedback (P. M. Lehrer & Gevirtz, 2014). It has been shown to promote a number of health benefits, including long-term emotional health (P. M. Lehrer & Gevirtz, 2014) and positive effects on cardiovascular diseases (Burlacu et al., 2021). During some meditative practices, breathing has been shown to slow, thus driving large heart rate oscillations at the breathing frequency (Bernardi et al., 2001; P. Lehrer et al., 1999; C.-K. Peng et al., 2004). For example, an expert meditator focusing on the breath during a Samadhi meditation showed high amplitude heart rate oscillations around 0.1 Hz (Phongsuphap et al., 2008), indicating that breathing slowed to around this frequency during one type of meditation. Similar increases in heart rate oscillations have been observed in Qigong, Kundalini yoga, or Zazen practitioners (P. Lehrer et al., 1999; C. K. Peng et al., 1999; C.-K. Peng et al., 2004) or in people reciting either the rosary or a typical yoga mantra (Bernardi et al., 2001). Tai Chi also increases abdominal breathing and heart rate oscillatory activity (Wei et al., 2016). However, not all meditative practices increase heart rate oscillations. For instance, in one study, the Vipassana Anapana practice as taught by S.N. Goenka, involving specific concentration on the minute sensations around the nostrils and the upper lip, does not increase high and low frequency HRV in long term daily practitioners (Delgado-Pastor et al., 2013). A recent systematic review (Karunarathne et al., 2024) found that of the five studies they found that assessed respiratory rate during meditation compared with baseline in long-term meditators, three studies — Vipassana meditators doing a breath awareness practice (Kodituwakku et al., 2012), Raja Yoga meditators during their practice (Sukhsohale & Phatak, 2012) and transcendental meditation practitioners during their practice (Wallace & Benson, 1972) — documented slower breathing during meditation practice than during baseline. Two other studies with long-term meditators doing an “OM” meditation (Telles et al., 1995; Telles & Desiraju, 1993) showed no changes in respiratory rate during their practice compared with baseline. Given the known health benefits of slow breathing, it is possible that meditative practices with or without slow breathing may lead to different types of health benefits, a hypothesis the current study set out to explore.

In our recent clinical trial (the Heart Rate Variability and Emotion Regulation or HRV-ER; Min et al., 2023), we found that randomizing participants to four weeks of daily practice of slow breathing reduced their plasma amyloid beta (Aβ) levels, levels of which are associated with increased risk for AD (F. Song et al., 2011), in both healthy younger and older adults (aged 18-35 and 55-80, respectively). In contrast, participants who were randomized to an active comparison group without slow paced breathing showed increases in plasma Aβ. We hypothesized that slow breathing decreases overall Aβ levels by stimulating an autonomic state that reduces Aβ production. Aβ is generated by two sequential cleavages of amyloid precursor protein (APP) by β and γ secretase (amyloidogenic pathway; right side of Fig. 1; Mather, 2025). However, if APP is first cleaved by α secretase rather than by β secretase, no Aβ is produced (nonamyloidogenic pathway; left side of Fig. 1). Cholinergic M1 and M3 receptors activate the α-secretase APP nonamyloidogenic pathway and suppress the β-secretase amyloidogenic pathway. Thus, it seems plausible that the slow breathing, which stimulates parasympathetic activity, concurrently promotes the nonamyloidogenic pathway, possibly through cholinergic M1 and M3 receptors, leading to a reduction of Aβ production. In contrast, noradrenergic receptor activity can bias APP cleavage towards the amyloidogenic pathway (Y. Chen et al., 2014; Ni et al., 2006). It is possible that the HRV-ER active control involving a meditative state while attending to heart rate biofeedback increased noradrenergic activity by focusing attention, and that noradrenergic activity biased APP processing to increase Aβ production.

**Fig. 1.**
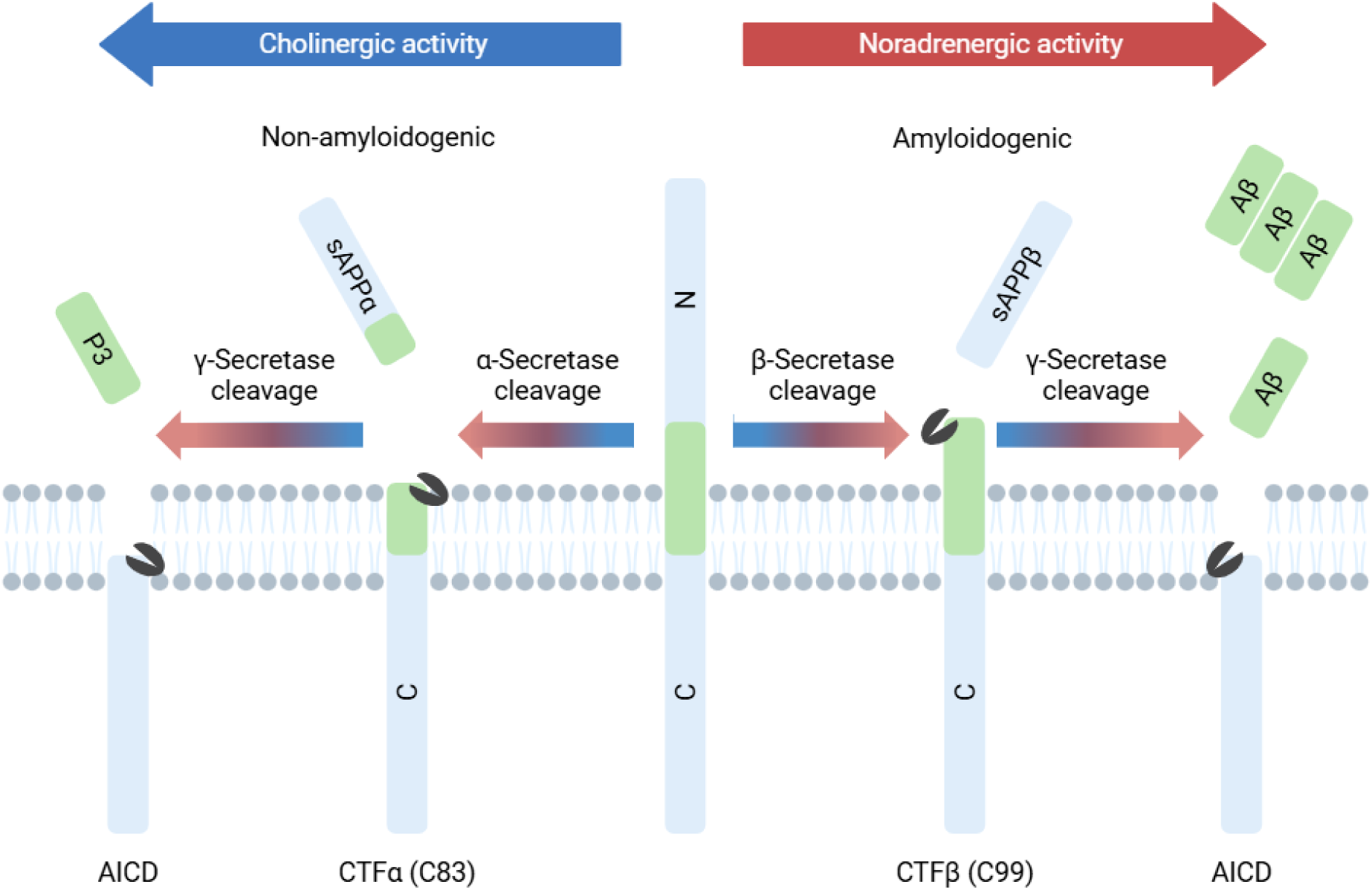
Cholinergic and Noradrenergic Activity Bias the Cleavage of Amyloid Precursor Protein (APP) to Produce Aβ or Not Note. Image adapted from Mather (2025).

According to the amyloid hypothesis (Hardy & Selkoe, 2002; Selkoe & Hardy, 2016), an imbalance between Αβ production and clearance initiates a cascade of events leading to progression of AD. Consistent with this hypothesis, Aβ42 levels in CSF start to increase in early adulthood and peak in middle adulthood (Luo et al., 2022). Plasma Aβ40 and Aβ42 levels are elevated in people with genetic risk of AD before onset of disease symptoms (Coppus et al., 2012; Ertekin-Taner et al., 2000, 2008; Head et al., 2011; Scheuner et al., 1996; Tokuda et al., 1997). Consistent with these higher plasma Aβ levels in at-risk individuals, a meta-analysis of longitudinal studies indicates that higher baseline Aβ40 and Aβ42 levels in cognitively normal individuals predict conversion to AD (F. Song et al., 2011).

While plasma Aβ levels are elevated during pre-symptomatic stages in older and other at-risk individuals, plasma Aβ42 levels decrease around the time of an AD diagnosis (F. Song et al., 2011). This results in decreased Aβ42 to Aβ40 ratios. High precision assays of plasma Aβ42/Aβ40 ratios correspond with amyloid PET (AUC 0.94; Schindler et al., 2019; F. Song et al., 2011; West et al., 2021). Aβ42 is more likely than Aβ40 to form aggregates in the brain (Masters & Selkoe, 2012); therefore, Aβ42 is less likely to be cleared from the brain to the periphery in individuals in whom amyloid deposition is actively occurring (Roberts et al., 2014). In contrast, Aβ42/Aβ40 ratios are not affected in younger adults who have not yet started to experience Aβ42 aggregation.

Our initial results showing that slow breathing practice reduced plasma Aβ levels raise an important question: Might slow breathing during meditation reduce plasma Aβ levels? As a first step, we examined in younger adults whether one week of daily mindfulness meditation practice with or without slow breathing instruction would differentially affect plasma Aβ levels and plasma Aβ42/Aβ40 ratios compared with no intervention controls. These were the primary outcome measures in our pre-registered parallel-group randomized clinical trial with a superiority framework (<u>ClinicalTrials.gov</u>; NCT06210035). We also tested if the two types of meditation practices would affect plasma pTau-181/tau ratio, another biomarker for brain clearance, and emotional well-being (secondary outcome measures). We hypothesized that 1) mindfulness meditation with slow breathing (M+SB) would reduce plasma Aβ40 and Aβ42 levels whereas mindfulness meditation without slow breathing (M-SB) and no intervention would not; 2) none of the three groups would show changes in plasma Aβ42/40 ratio or plasma pTau-181/tau ratio since participants in this study are too young (18-35 years) to have brain Aβ and tau deposition; 3) both meditation conditions (M+SB and M-SB) would show greater improvement in emotional well-being compared with the no-intervention condition. Although it was not the focus of the study, we also tested the hypothesis that the M+SB condition would show greater episodic memory for positive items relative to the other conditions (pre-registered as other pre-specified outcome measures), as indicated in our preliminary findings (Cho et al., 2023).

## Methods

### Pilot Studies

Before running the actual study, we conducted three pilot experiments to identify two types of mindfulness meditation practices to compare in our study: one incorporating slow breathing and the other without it. Previous research found that “anapanasati” (mindfulness of breath) practice in long term Vipassana meditators in the Goenka tradition (with a nostril focus) did not increase LF-HRV compared with rest or random thinking (Delgado-Pastor et al., 2013), suggesting that mindfulness of breath practice may not necessarily elicit slow breathing. We thought that this might have to do with the location of the focus of attention. Participants in this study focused on the nostrils where the passage of air can make it harder to detect minute sensation, which might have quieted their breathing (shallow breathing rather than slow breathing). In contrast, we thought that the more typical breath-focused mindfulness that directs attention to the belly would encourage slow breathing. We pilot tested this comparison (belly-focus, nostril-focus or control; see Supplementary Table 1 for the meditation scripts) with 6 participants doing the 20-minute practice twice a day for 7 days while recording their pulse (two people in each condition). The control group was instructed to do whatever they wanted as long as their activity did not involve motion (e.g., use your phone or computer, read a book). To our surprise, we found that neither meditation condition increased LF-HRV much relative to control (Supplementary Figure 1a), indicating that neither elicited slowed breathing (we had expected the belly-focus condition to slow down breathing). In the second pilot study, we tried modifying the instructions to make breathing differ more between the two meditation conditions with another 6 people (this time just practicing once for 20 minutes). As shown in Supplementary Table 2, we attempted to encourage the belly-focus group to breathe slowly and guide the nostril-focus group to breathe quietly without explicitly instructing them to change their breathing rate. However, we found that neither group increased LF-HRV, indicating no changes in breathing rate, relative to their own resting state (Supplementary Figure 1b). In the third pilot study, we provided explicit instructions to breathe slowly to one meditation group but not the other (Supplementary Table 3). Since we thought that it is also important to keep the two meditation conditions the same except for the presence or absence of slow breathing, we decided to give the same meditation instructions (i.e., belly-focus) to both groups. Three participants performed both types of meditations for 10 min each in a random order as well as a 5-min rest. The belly-focus mindfulness meditation with the slow breathing instructions increased LF-HRV relative to rest, which was not observed in the belly-focus meditation without slow breathing instructions (Supplementary Figure 1c). The three pilot experiments informed us that slow breathing does not naturally occur in novice meditators during mindful attention to breath, and therefore we needed to provide explicit instructions to incorporate slow breathing during meditation. The final design of this study is described in the “Overview of 1-week Protocol Schedule” section below.

### Participants

We recruited 94 participants from online advertisements (e.g., University of Southern California [USC] Healthy Minds Research Volunteer Registry, USC Slack student groups, Volunteer Match), flyers or personal referrals. Participants provided informed consent approved by the USC Institutional Review Board. Participants were screened to ensure that they speak English fluently, are between age 18 and 35, agree to provide blood samples and devote 40 minutes daily for an assigned intervention, and did not currently practice any relaxation, biofeedback, or breathing technique (e.g., meditation) for more than an hour a week. We excluded people with abnormal cardiac rhythms, dyspnea, or heart disease. Two people dropped out of the study before the intervention began (one participant fainted right after the blood draw and another participant was no longer interested in participating in the study). Three people withdrew from the study after the intervention began since they could not commit to complete home practice as they originally thought. The remaining 89 participants were included in our analyses (Fig. 2). Age, sex and education were similar in the three conditions (Table 1).

**Fig. 2.**
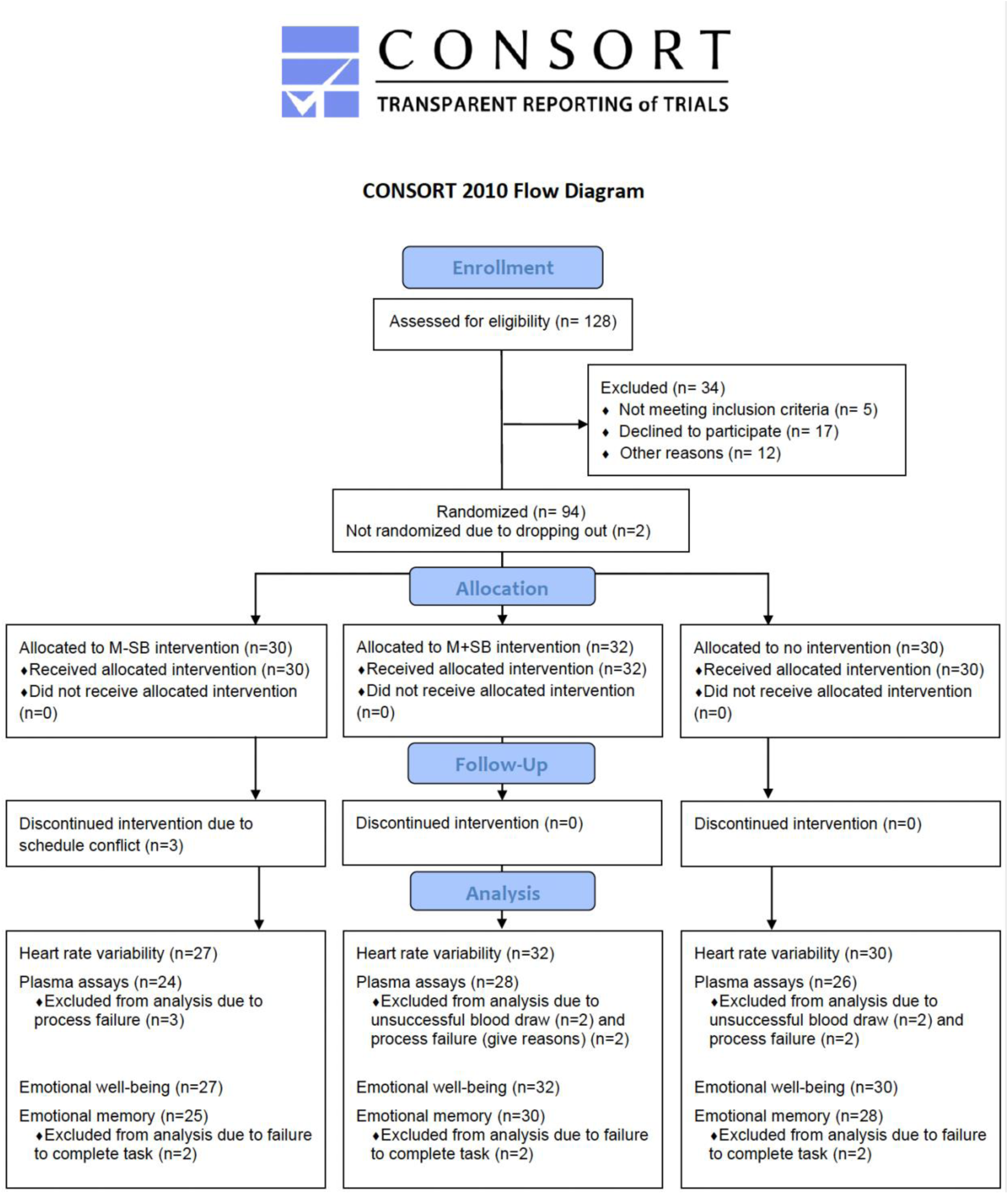
CONSORT Flow Diagram *Note.* Number of participants who enrolled, were allocated to conditions and included in each analysis.

**Table 1.** Characteristics of the Study Participants.

|  | **Age** |  | **Years of education** |  | **Sex** |  |
| --- | --- | --- | --- | --- | --- | --- |
|  | *Mean (SD)* | *Range* | *Mean (SD)* | *Range* | *Female* | *Male* |
| Belly (n=27) | 25.07 (SD=5.16) | 19-35 | 15.88 (SD=1.86) | 12-19 | 21 | 6 |
| Belly + SB (n=32) | 26.25 (SD=5.29) | 18-35 | 16.19 (SD=1.93) | 12-20 | 26 | 6 |
| No Intervention (n=30) | 26.20 (SD=4.82) | 18-35 | 16.47 (SD=2.56) | 13-23 | 25 | 5 |

### Overview of 1-week Protocol Schedule

Participants visited the USC Emotion and Cognition lab twice, once before and once after the one-week intervention. During both visits, participants completed a blood draw, questionnaires (see the *Questionnaires* section for more detail) and a seated resting pulse measure for 5 minutes. During the 5-min resting state, participants were instructed to sit in a chair with palms up, both feet flat on the floor, back straight, knees 90-degree angle, and eyes closed. Participants wore an ear sensor to measure their pulse, which was connected to a USB module plugged into a USB port on the training laptop computer. The pulse data was recorded by emWave pro software and an associated sensor unit (Heartmath, 2016). Data were saved in a database file on the laptop. In addition, participants performed 5 minutes of paced breathing at 15 breaths per minute by following a visual pacer, which allowed us to measure HRV while controlling for breathing pace. The customized application randomly assigned participants to one of the three conditions (belly-focus concentration meditation, belly-focus concentration meditation with slow breathing, no-intervention control), stratified by participant sex with a block size of three. Between the two lab visits, participants were instructed to engage in the assigned practice for 20 minutes, twice a day (i.e., a total of 40 minutes/day). The first 20-minute practice session was performed during the first visit under the experimenter’s supervision. The rest of the practice was conducted at home.

### Mindfulness Meditation without Slow Breathing Condition (M-SB)

During home practice, participants used the same ear sensor, USB cable and laptop as used in the lab. Participants viewed the instructions on the customized app while the emWave pro software (Heartmath, 2016) ran in the background, which was not visible to the participants. The app showed instructions, “You will be asked to sit quietly to meditate for 20 minutes while your heart rate is recorded. During this time, you will listen to a guided meditation to help you focus on the sensations around the belly. Please remember that it is normal for the mind to wander. When this happens, gently bring your attention back to your belly.” Once participants pressed start, the recording began. Participants were asked to gently close their eyes and focus their attention on the sensations around the belly. The entire guided meditation script can be found in Supplementary Table 4.

### Mindfulness Meditation with Slow Breathing Condition (M+SB)

Participants were given the same devices and instructions as in the belly-focus meditation condition except that the app included an additional instruction to breathe slowly during the entire meditation session. Once participants pressed start, the recording began and asked participants to gently close their eyes. Participants were guided to focus their attention on the sensations around the belly while breathing slowly at around 10 seconds per breath. The additional instruction provided in this condition by the guiding voice was, “Take long slow breaths as I guide you. Inhale for a count to 5 and exhale for a count to 5. Breathe slowly and gently at this pace during the entire meditation exercise. I’ll count with you for the first few breaths. Then you can count on your own. Inhale, 2, 3, 4, 5, exhale 2, 3, 4, 5. Inhale, 2, 3, 4, 5, exhale 2, 3, 4, 5. Continue breathing this way on your own.” The entire guided meditation for this condition can be found in Supplementary Table 4.

### No-intervention Control Condition

Participants were given the same devices as in the two other conditions. The app instructed, “You will be asked to sit quietly for 20 minutes while your heart rate is recorded. Please feel free to do whatever you would like to do (e.g., use your phone or computer, read a book, watch TV, listen to music) as long as your activity does not involve head motion.” Once participants pressed start, the recording began as participants started the activity of their choice.

### Coaching

During the initial 20-minute practice session in the lab, the experimenter monitored the participant’s real time heart rate data to ensure that their breathing patterns aligned with their conditions (i.e., M+SB participants who were instructed to breathe slowly around 10s/breath were expected to show large heart rate oscillations whereas M-SB participants were expected to breathe naturally as reflected by small heart rate oscillations). Four participants in the M+SB condition failed to breathe slowly. Two participants in the M-SB condition failed to breathe naturally (they breathed slowly). We stopped their sessions before reaching the half-way point (i.e., 10 minutes), instructed them to breathe according to their condition, and restarted their 20-min session. Since this additional coaching may have disrupted their natural physiological state during meditation, we repeated all the data analyses reported in the results section excluding these six participants. However, the results remain the same. The time-by-condition interactions for Aβ42 and Aβ40 remained significant. The rest of the results were non-significant.

### Blood Collection Procedure

The blood samples were collected via antecubital venipuncture between 9:00AM and 5:30PM. The time of the day for blood collection was consistent across both visits within each participant. A phlebotomist drew 10 ml of blood from each participant’s arm into a K2 EDTA tube. To separate plasma from red blood cells, the whole blood from the K2 EDTA tubes was centrifuged at the speed of 1500 RPM for 15 min at room temperature (15°C). Plasma was aliquoted in cryovials and stored at −80 °C. The frozen plasma samples were transferred and assayed at University of California, Irvine for Aβ42, Aβ40, total tau (tTau), and phosphorylated tau (pTau-181).

### Questionnaires

Participants completed a demographic questionnaire during their first visit. At pre- and post-intervention lab visits, they completed the profile of mood states (Grove & Prapavessis, n.d.), the sort-form version of depression anxiety stress scales (Lovibond & Lovibond, 1995) and the 15-item version of the five facet mindfulness questionnaire (Baer & Carmody, 2012; Gu et al., 2016). We used the 40-item version of POMS. Participants indicated how much each item reflected how they felt at the moment using a scale from 1 (not at all) to 5 (extremely). Total mood disturbance was calculated by subtracting positive-item totals from negative-item totals. A constant value (i.e., 100) was added to the total mood disturbance to eliminate negative scores. Higher scores indicate greater negative affect. The DASS-21 consisted of 21 items with three subscales: depression, anxiety, and stress. Each subscale comprises seven items, which reflect a symptom of depression, anxiety, and stress. Participants reported how much each statement applied to them in the past week using a 4-point scale ranging from 0 (did not apply to me at all) to 3 (applied to me very much or most of the time). The total points for each category were summed up and multiplied by 2. Higher scores indicate more frequent symptomatology. The FFMQ consisted of five subscales: observing, describing, acting with awareness, nonjudging of inner experience and nonreacting of inner experience. Participants responded to each item using a 5-point scale ranging from 1 (never/rarely true) to 5 (very often/always true). We reverse-scored negatively worded items and summed items to create subscale and total scores. Since it is recommended that researchers should exclude the observing facet score from comparisons of total scale/subscale scores before and after mindfulness interventions (Gu et al., 2016), we excluded the observing score from our analyses.

### Emotional Memory Task

We measured episodic memory as a pre-specified outcome measure (not primary or secondary) since we were interested in the effects of the interventions on emotional memory. Before each 20-minute practice session, the customized application showed participants six images, two from each valence category (positive, negative, neutral) from the Nencki Affective Picture System (NAPS; Marchewka et al., 2014) using jsPsych (de Leeuw et al., 2023). They were asked to rate each picture on a scale of 1-9 (1 = very negative to 9 = very positive, with 5 = neutral). At the post-intervention visit, they were given surprise recall and recognition memory tests on those images. First, participants were given a free recall test where they were asked to describe in detail as many images as they could that they saw in the past week. There was no time limit imposed for the free recall. Next, participants were given a recognition test where they viewed the 84 old images and 42 previously new images. For 16 people who did not encode all pictures since they missed some practice sessions, we adjusted the total number of pictures when calculated their memory scores (e.g., if participants missed 2 practices, which means they missed to encode 12 pictures, we subtracted 12 from 84 old images).

### Analyses

For heart rate variability analyses, we conducted a 2 (session type: baseline vs. training) x 3 (condition: M+SB, M-SB, no intervention) ANOVA. We performed 2 (time: pre-, post-intervention) x 3 (condition: M+SB, M-SB, no intervention) ANOVAs to test the intervention effect on all outcome measures unless otherwise mentioned. Post-hoc paired samples t-tests were conducted to compare pre- and post-intervention values for each condition.

#### Heart Rate Variability during Home Practice and Seated Rest

The pulse data recorded by emWave pro software and sensor unit were saved in a database file on the laptop. The database file was also transferred to a remote server via internet connection by custom software. Interbeat interval (IBI) data were exported from the database file for HRV analysis. HRV data were analyzed using Kubios HRV Premium 3.1 (Tarvainen et al., 2014) with the following parameters: artifact correction = automatic, smoothing prior lambda = 500, AR model order = 16. Among the HRV indices, we used autoregressive spectral power in low frequency (LF 0.04-0.15 Hz) and high frequency (HF 0.15-0.4 Hz) bands, root mean squared successive difference (RMSSD) and mean heart rate for each practice session and for 5-minute rest sessions during pre- and post-intervention lab visits. For each participant, **all HRV indices** from practice sessions were averaged across sessions. We excluded data with extreme values of RMSSD (> 200), which were 18 training sessions and 1 post-intervention resting session. Before conducting statistical analyses, we log transformed the LF and HF power values due to non-normality.

#### Plasma Assay Procedure

Plasma samples were analyzed using the automated Simoa SR-X analyzer with the commercially available Simoa Human Neurology 3-Plex A assay kit (Quanterix, Billerica, MA, USA) for Aβ42, Aβ40, and tTau. Plasma concentrations of pTau-181 were measured using the automated Simoa HD-X analyzer and the Simoa pTau-181 Advantage V2 kit (Quanterix, Billerica, MA, USA). Analyses were performed in duplicates (mean % coefficient of variation [%CV]: Aβ42: 6.33, Aβ40: 4.55, tTau: 8.81, pTau-181: 6.01) using a 1:4 dilution protocol according to the manufacturer’s instructions. Prior to analysis, plasma samples were thawed at room temperature for 1 h and centrifuged at 10,000×g for 5 min to prevent transfer of debris. All assays were conducted by the same operator with the same respective instrument. As in our previous study (Min et al., 2023), we defined outliers who on a box-and-whisker plot were above Q3 + 3 * the interquartile range on each of plasma measures (Aβ42, Aβ40, Aβ42/Aβ40 ratio, pTau/tTau ratio, pTau-181, tTau) for pre- and post-intervention separately. This resulted in excluding outliers for pre-intervention Aβ42/Aβ40 ratio (N=3), post-intervention Aβ42/Aβ40 ratio (N=1), pre-intervention pTau (N=1), pre-intervention tTau ratio (N=4), and post-intervention tTau ratio (N=2), pre-intervention pTau/tTau ratio (N=4), and post-intervention pTau/tTau ratio (N=3).

We examined the relationships between pre-to-post changes in plasma biomarkers and HRV during practice. We also examined the correlations between changes in plasma biomarkers and changes in HRV. For these correlation analyses, a significant threshold was set at false discovery rate (FDR) *p* < 0.05.

#### Emotional Well-being

We defined outliers for each emotion measure if their pre-intervention or post-intervention values were greater than the third quartile plus three times of interquartile range or lower than the first quartile minus three times of the interquartile range based on the boxplot rule of SPSS. This resulted in excluding outliers for pre-intervention POMS (N=4), post-intervention depression (N=2), and pre-intervention FFMQ (N=1).

#### Emotional Memory

Following the method used in our previous study (Cho et al., 2023), we created a summary positive emotional memory bias score with false recognition and recall contributing equally to the formula. The *Z* scores for positive and negative false alarms and correct recall counts were calculated across participants. This means that the average false alarm rates and correct recall counts for positive and negative images were set to zero and variability was normalized such that scores of ± 1 reflected a bias one standard deviation from the mean for that type of item. We computed positive emotional memory bias score with the following formula: (Z FalseAlarmPositive—Z FalseAlarmNegative) + (Z CorrectRecallPositive—Z CorrectRecallNegative). We performed a one-way ANOVA to assess condition differences on the positive emotional bias score. In addition, we conducted separate analyses for recall and recognition performance (see Supplementary Information).

## Results

### Heart Rate Oscillations during Training vs. Rest

We first examined whether M+SB participants breathed slowly during practice as they were instructed, which should lead to higher heart rate oscillation power in the low frequency range (0.04-0.15 Hz or 7-25 sec) compared with their own baseline rest. Slow breathing was not anticipated in the M-SB and no-intervention conditions. As expected, we found a significant interaction of session type (baseline vs. training) and condition (M+SB, M-SB, no intervention), *F*(1,87) = 28.98, *p* < 0.001, *η*_p_ ^2^ = .40. M+SB participants increased LF power during practice compared with rest, *t*(30) = 7.62, *p* < 0.01, while such a difference was not observed in M-SB, *t*(28) = 1.23, *p* = 0.231, and no intervention, *t*(29) = -1.46, *p* = 0.154 (Fig. 3). Other HRV parameters during practice, pre- and post-rest, and controlled breathing (e.g., LF, HF, RMSSD, heart rate) are reported in Supplementary Tables 5-7.

**Fig. 3.**
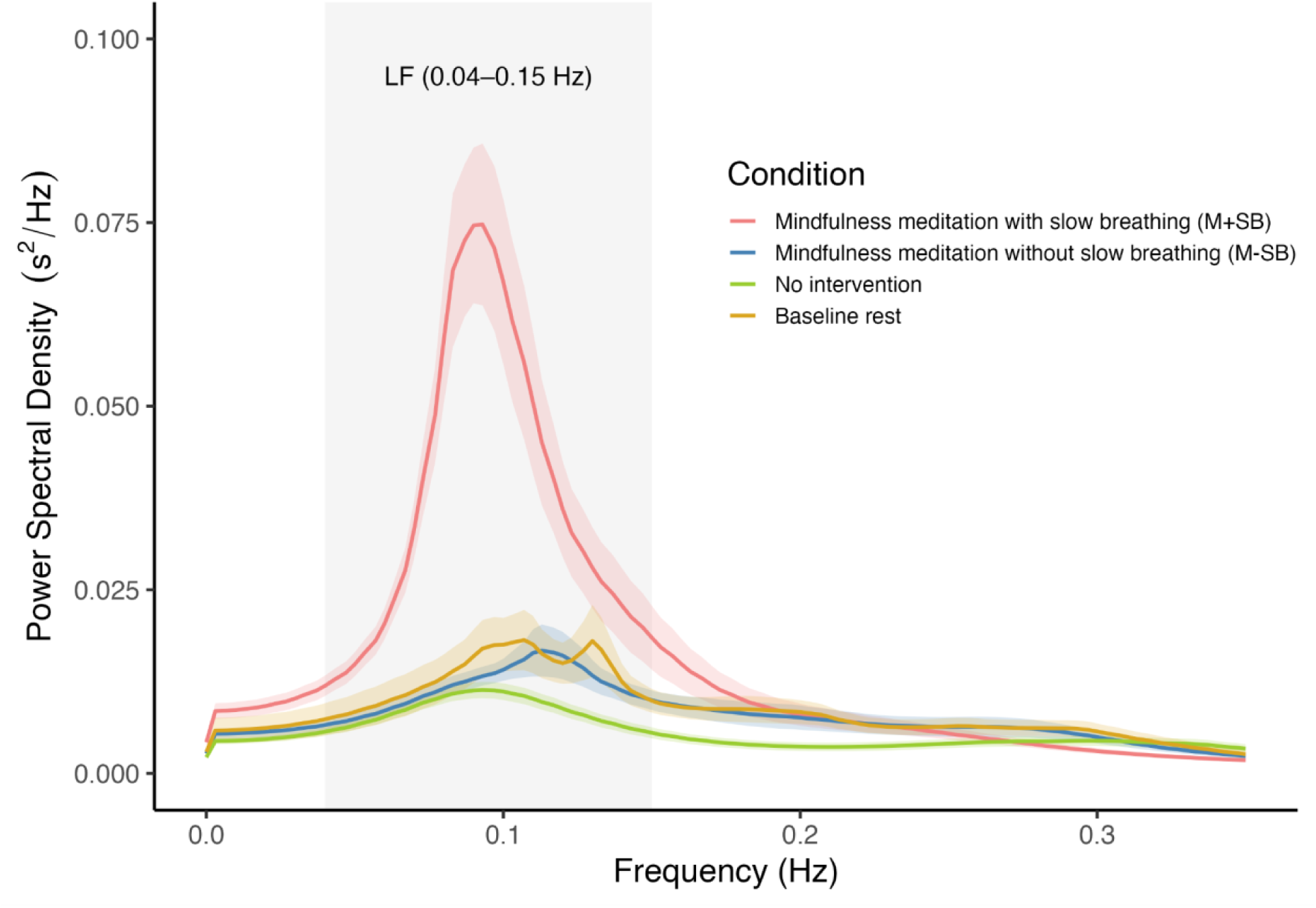
Comparisons of Heart Rate Power Spectral Density in the Three Conditions *Note.* Error bands represent the standard errors. Power spectral density for baseline rest is the average across the three conditions.

### Intervention Effects on Plasma Aβ42, Aβ40 and Aβ42/Aβ40 Ratio (Primary Outcome Measures)

There was a significant 2 (time: pre, post) by 3 (condition: M+SB, M-SB, no intervention) interaction for an aggregate score between Aβ42 and Aβ40 levels, *F*(2, 75) = 6.21, *p* = 0.003, *η*_p_ ^2^ = 0.14. The interaction was driven by M+SB reducing Aβ levels from pre- to post-intervention, *t*(27) = -2.67, *p* = 0.013, M-SB increasing Aβ levels, *t*(23) = 2.53, *p* = 0.019, and no-intervention showing no changes, *t*(23) = -0.20, *p* = 0.847. When we examined Aβ42 and Aβ40 levels separately, there were also significant time-by-condition interactions for Aβ42, *F*(2, 75) = 6.86, *p* = 0.002, *η*_p_^2^ = 0.16 , and for Aβ40, *F*(2, 75) = 3.87, *p* = 0.025, *η*_p_^2^ = 0.09 (Fig. 4a-4b). Aβ42 levels were decreased in M+SB, *t*(27) = -2.10, *p* = 0.045, increased in M-SB, *t*(23) = 3.31, *p* = 0.003, and showed no changes in no intervention, *t*(25) = 0.09, *p* = 0.993. Aβ40 levels were increased in M-SB, *t*(23) = 2.47, *p* = 0.021, and showed no changes in M+SB, *t*(27) = -1.51, *p* = 0.142 (with a decreasing trend), and no intervention, *t*(25) = 0.38, *p* = 0.707. As expected, we did not find a significant time-by-condition interaction for Aβ42/Aβ40 ratio, *F*(2, 72) = 0.78, *p* = 0.460, *η*_p_^2^ = 0.021. The means of all biomarkers for each condition and for each time point are reported in Table 2.

**Fig. 4.**
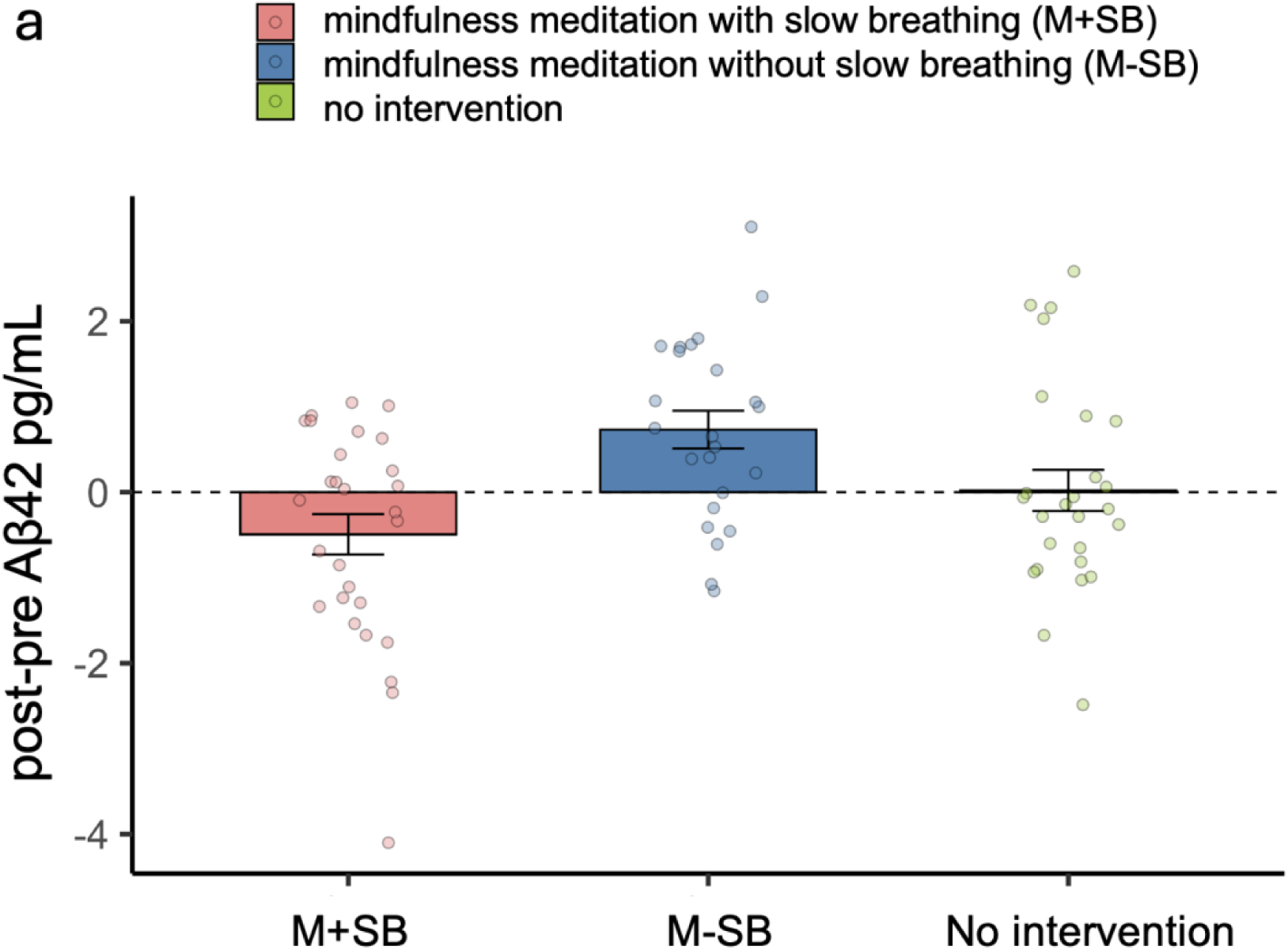
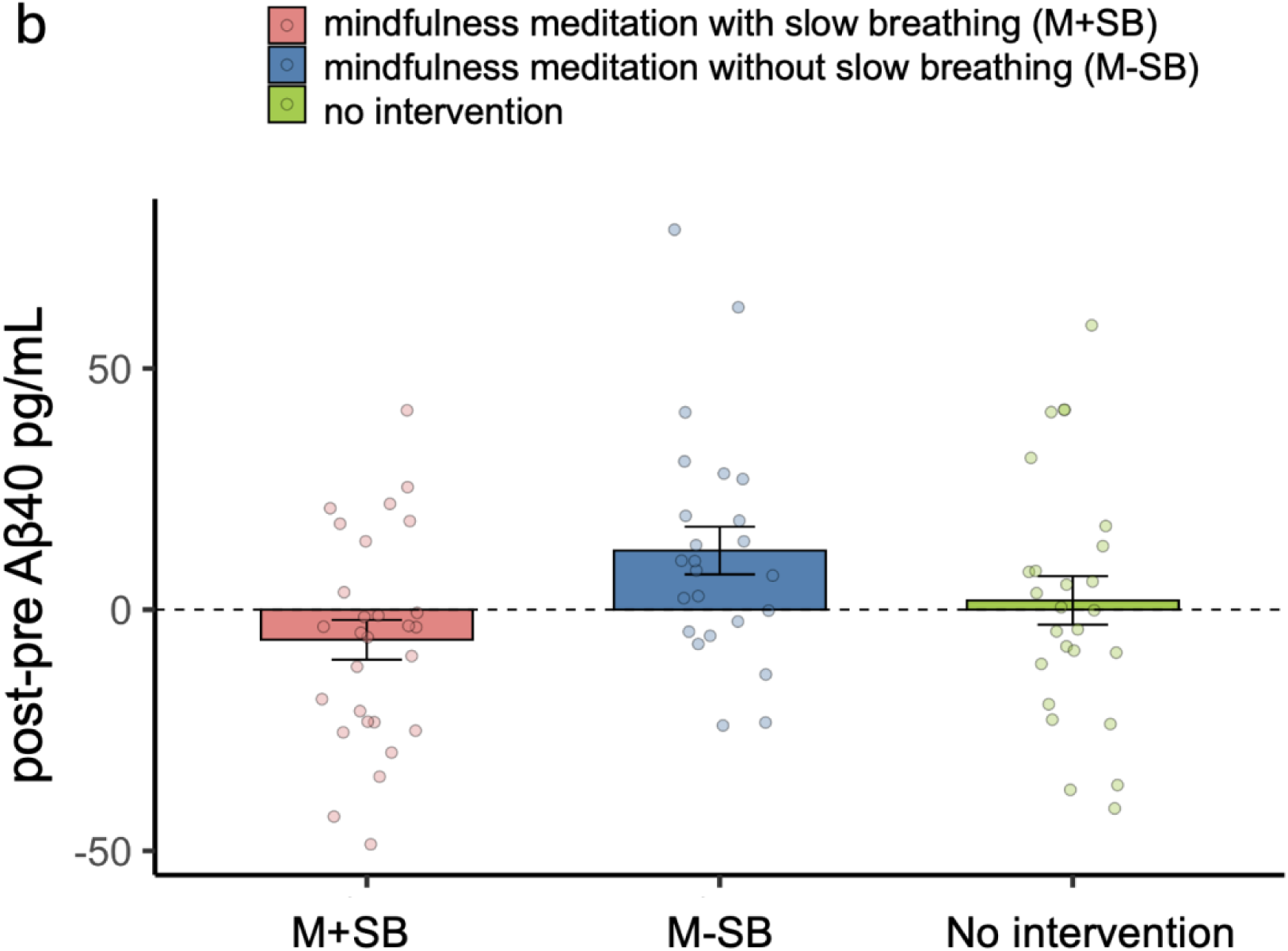
Pre-to-post Changes in Plasma Aβ42 (a) and Aβ40 (b) *Note.* One week of daily mindfulness meditation with (red bars) and without (blue bars) slow-paced breathing had opposite effects on plasma Aβ levels.

**Table 2.**
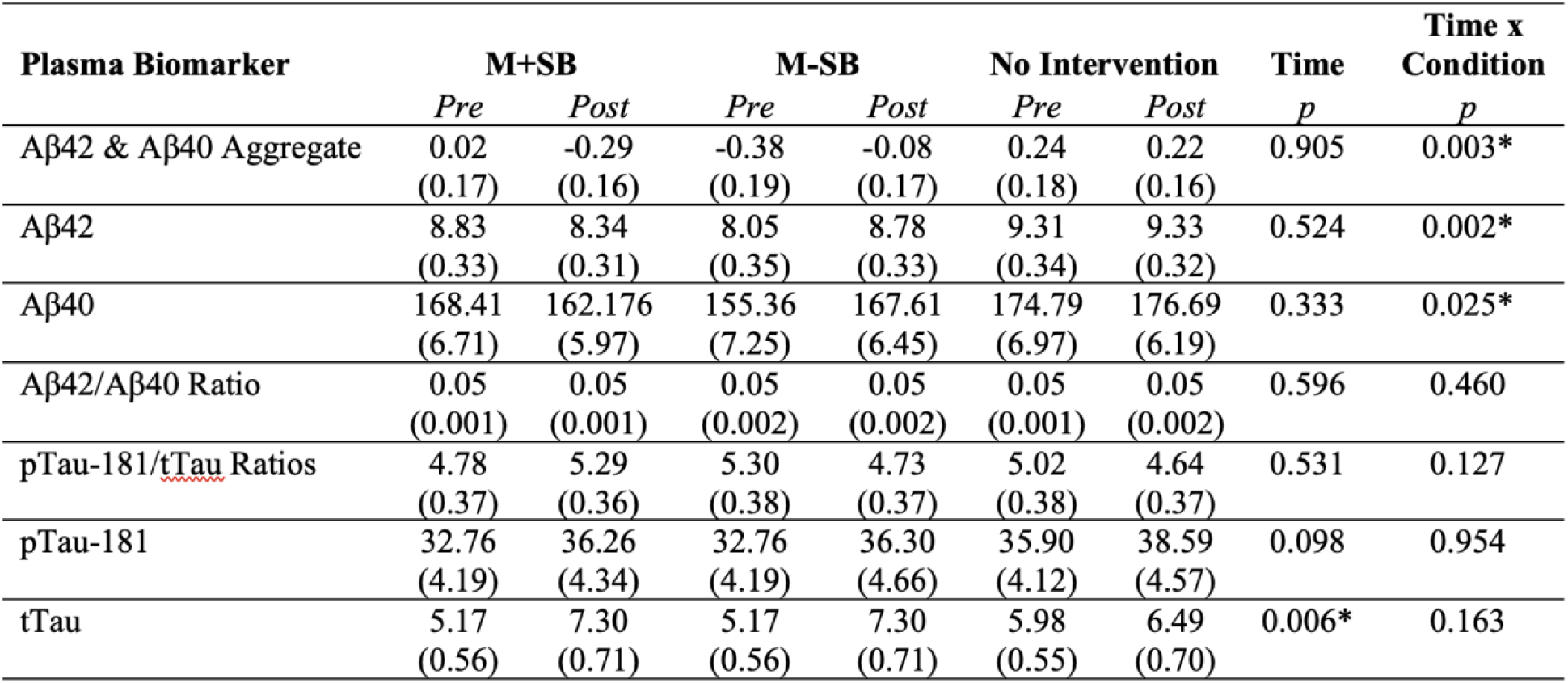
Means of All Biomarkers for Each Condition at Pre- and Post-intervention.

| Plasma Biomarker | M+SB |  | M-SB |  | No Intervention |  | Time<br>*p* | Time x<br>Condition<br>*p* |
| --- | --- | --- | --- | --- | --- | --- | --- | --- |
|  | *Pre* | *Post* | *Pre* | *Post* | *Pre* | *Post* |  |  |
| Aβ42 & Aβ40 Aggregate | 0.02<br>(0.17) | -0.29<br>(0.16) | -0.38<br>(0.19) | -0.08<br>(0.17) | 0.24<br>(0.18) | 0.22<br>(0.16) | 0.905 | 0.003* |
| Aβ42 | 8.83<br>(0.33) | 8.34<br>(0.31) | 8.05<br>(0.35) | 8.78<br>(0.33) | 9.31<br>(0.34) | 9.33<br>(0.32) | 0.524 | 0.002* |
| Aβ40 | 168.41<br>(6.71) | 162.176<br>(5.97) | 155.36<br>(7.25) | 167.61<br>(6.45) | 174.79<br>(6.97) | 176.69<br>(6.19) | 0.333 | 0.025* |
| Aβ42/Aβ40 Ratio | 0.05<br>(0.001) | 0.05<br>(0.001) | 0.05<br>(0.002) | 0.05<br>(0.002) | 0.05<br>(0.001) | 0.05<br>(0.002) | 0.596 | 0.460 |
| pTau-181/tTau Ratios | 4.78<br>(0.37) | 5.29<br>(0.36) | 5.30<br>(0.38) | 4.73<br>(0.37) | 5.02<br>(0.38) | 4.64<br>(0.37) | 0.531 | 0.127 |
| pTau-181 | 32.76<br>(4.19) | 36.26<br>(4.34) | 32.76<br>(4.19) | 36.30<br>(4.66) | 35.90<br>(4.12) | 38.59<br>(4.57) | 0.098 | 0.954 |
| tTau | 5.17<br>(0.56) | 7.30<br>(0.71) | 5.17<br>(0.56) | 7.30<br>(0.71) | 5.98<br>(0.55) | 6.49<br>(0.70) | 0.006* | 0.163 |

### Intervention Effects on Plasma pTau-181/tTau Ratios (Secondary Outcome Measures)

As hypothesized, there was no significant time-by-condition interaction for pTau-181/tTau ratio, *F*(2, 68) = 2.13, *p* = 0.127, *η*_p_ ^2^ = 0.059 (the means are reported in Table 2). Also there was no significant time-by-condition interaction for pTau-181, *F*(2, 80) = 0.05, *p* = 0.954, *η*_p_ ^2^ = 0.001, or for tTau, *F*(2, 71) = 1.86, *p* = 0.163, *η*_p_ ^2^ = 0.050.

### Intervention Effects on Emotional Well-being (Secondary Outcome Measures)

Since there were significant baseline differences in several emotion measures between the conditions (i.e., mood, depression and mindfulness; the means are reported in Supplementary Table 8), we performed univariate analyses comparing post-intervention scores of the three conditions while controlling for pre-intervention scores of each emotion outcome variable. However, there were no significant condition differences in any of these measures at post intervention (Supplementary Table 8), suggesting that neither meditation intervention had a significant impact on participants’ subjective mood, stress, anxiety, depression or mindfulness. Two-way ANOVA results are also reported in the “Intervention effects on emotional well-being” section of Supplementary Information.

### Intervention Effects on Emotional Memory (Pre-registered as Other Pre-specified Outcome Measure)

Since our preliminary findings suggest that a slow-paced breathing intervention influences positive emotional memory (Cho et al., 2023), we first calculated a summary positive emotional memory bias score with false recognition and recall contributing equally to the formula (see the Analyses section in Methods for more details). There was no significant condition difference in positive emotional memory bias score, *F*(2, 80) = 0.424, *p* = 0.656, *η*_p_^2^ = 0.010. We also examined condition differences in recall and recognition performances separately, but no significant results were found (see “Intervention effects on emotional memory” in Supplementary Information). Additional Analyses on the Relationships between HRV and Plasma Biomarkers

We conducted exploratory correlation analyses between HRV during practice and pre-to-post changes in plasma biomarkers (Supplementary Table 9) and between pre-to-post changes in HRV and those in plasma biomarkers (Supplementary Table 10), but no significant correlations were found.

## Discussion

This study examined whether a short course of one week of practicing mindfulness meditation with or without slow-paced breathing would differentially affect plasma biomarkers for AD and emotional well-being. We found support for our first hypothesis that meditative practice incorporating slow breathing would reduce plasma Aβ levels whereas the other conditions would not. We also found support for our second hypothesis that plasma Aβ42/40 ratio and plasma pTau-181/tau ratio would not be affected by any of the interventions. We did not find support for our third hypothesis that both meditation conditions would improve emotional well-being relative to a no intervention control.

As discussed earlier, slow breathing may affect Aβ levels by modulating Aβ production. Our hypothesis is that slow breathing drives the α-secretase APP nonamyloidogenic pathway and suppresses the β-secretase amyloidogenic pathway, leading to a reduction of Aβ production. This may explain the Aβ reduction observed in the M+SB condition. The Aβ increase in the M-SB condition is intriguing, as it is consistent with the ‘Osc-’ condition in our prior HRV-ER clinical trial, who also showed increases in plasma Aβ (Min et al., 2023). Like the current M-SB condition, the Osc-condition was not doing slow breathing (they received heart rate variability biofeedback that guided them to avoid heart rate oscillations) and the study instructions suggested they should attempt to meditate. One possible explanation is that attempting to meditate might have required cognitive effort in these novice meditators, which in turn may have led to elevated arousal/noradrenaline levels leading to increased plasma Aβ. While both meditation groups were instructed to focus their attention on the sensations around the belly, M+SB participants might have been able to counteract increased levels of arousal due to the focused attention task by parasympathetic (vagus) stimulation via the slow breathing. In more naturalistic common mindfulness meditation breath-focused practice instruction includes the explicit encouragement to allow the body to relax and/or taking a few deep breaths at the outset to encourage relaxation. The fact that these instructions were not included may also contribute to the effects seen in the M-SB group as the instructions were focused more purely on the attentional aspect without emphasizing the relaxation aspect of practice.

Another element of this study worth highlighting is that changes in plasma Aβ levels were observed after a one-week intervention, which was much shorter than the 4-week slow breathing intervention in our previous study (Min et al., 2023). Our results indicated that the effects of slow-paced breathing on plasma Aβ can occur relatively quickly, which is consistent with prior findings of overnight changes in plasma Aβ levels (Lucey et al., 2018) or even within hours (Lucey et al., 2018; Ovod et al., 2017).

None of the three conditions affected plasma Aβ42/40 ratios and plasma pTau-181/tau ratio. As discussed earlier, plasma Aβ42 levels decrease around the time of an AD diagnosis (F. Song et al., 2011), resulting in a reduction in plasma Aβ42/Aβ40 ratios. Aβ42 is more likely than Aβ40 to form aggregates in the brain (Masters & Selkoe, 2012), and less likely to be cleared from the brain to the periphery in those in whom amyloid deposition is actively occurring (Roberts et al., 2014). Consistent with this idea, our younger participants who presumably have not started to experience Aβ42 aggregation did not show changes in plasma Aβ42/Aβ40 ratios. The result was consistent with our prior findings that slow-paced breathing increased plasma Aβ42/Aβ40 ratios in older adults but not in younger adults (Min et al., 2023).

In contrast with physiological changes, the psychological benefits of meditation may take time to manifest. We failed to observe improvements in emotional well-being, including mood, stress, anxiety, depression and mindfulness. We also did not observe condition differences in positivity bias in episodic memory. The lack of behavioral changes may partially be due to limitations in the study design. It is possible that the duration of intervention was too short, as some prior findings suggest that longer practice was associated with greater behavioral benefits (Basso et al., 2019; Fredrickson et al., 2017; Lacaille et al., 2018). Another possibility is that participants needed more guidance to navigate through the initial difficulty in learning meditation. Future studies should incorporate these considerations.

To our knowledge, our initial findings on the effects of slow breathing intervention on plasma Aβ levels (Min et al., 2023) are the first from a randomized clinical trial to show that a behavioral intervention can reduce Aβ levels (measured via plasma, CSF or PET imaging) in healthy adults relative to a control group. The current study further sheds light on how slow breathing in the context of mindfulness meditation could help reduce plasma Aβ levels. In the current and previous studies, our outcome measure was plasma Αβ. Thus, a critical next step is to investigate what happens to Αβ42 levels in the brain. Plasma Αβ reflects both peripheral and brain sources of Αβ, and slow breathing may or may not have similar effects on Αβ in the brain as in the periphery. Future studies should examine whether this intervention has similar effects on Aβ levels in the periphery (blood) and in the brain (CSF). It is also important to extend this study to middle-aged and older adults who may be starting or have already started to have Aβ42 aggregate in the brain.

Future studies using fMRI would be informative for understanding common and separate neural mechanisms underlying M+SB vs. M-SB. A meta-analysis (Ganesan et al., 2022) suggests that focused attention meditation (which is similar to the one used in this study) involves altering the dynamics of the default mode network (DMN), the salience network and the executive control network. Dysfunction of the same networks with an additional involvement of the locus coeruleus (LC) have been implicated in age-related attentional deficits and distractibility (Lee et al., 2020). The LC is the brain’s primary source of noradrenaline and involved in arousal, attention, memory, respiration and the neuropathological progression of AD (Dahl et al., 2019, 2020, 2022; Gargaglioni et al., 2010; Mather & Harley, 2016; Yackle et al., 2017). One possibility worth pursuing in future research is that phasic stimulation of LC via the M+SB intervention may enhance the functional organization of the LC-networks, which may in turn lead to behavioral improvements, such as reduced distractibility and enhanced cognitive performance. On the other hand, the M-SB intervention may affect these networks differently.

From a practical perspective, these findings contribute to a body of evidence suggesting that some of the benefits of meditative practice may be related to slowing of the breath (Karunarathne et al., 2023, 2024; Kral et al., 2023; Wielgosz et al., 2016). Given this, studies assessing the relative benefit of arriving at slow breathing rates without intentional effort through the relaxation that occurs from engaging in practice regularly over time vs. as a result of purposeful slowing of the breath may be justified. Mainstream schools of mindfulness meditation practice often do not instruct students to engage in slow breathing out of a sense that controlling the breath might lead away from the experience of equanimity and acceptance - accepting the breath exactly as it naturally occurs. More rigorous exploration of the pros and cons of including slow breathing instructions in meditative interventions could help identify optimal ways to teach meditation to different populations depending on their baseline characteristics and desired outcomes. On the other hand, focused attention meditative practices that do not involve slowing of the breath may have other benefits, such as improving attention performance even when not meditating (Sumantry & Stewart, 2021; Verhaeghen, 2021).

This study demonstrated that physiological aspects of meditation can influence plasma biomarkers associated with AD. Our findings also highlight that this area of research deserves more attention and further investigation. Future studies should examine the underlying mechanisms of the effects of slow breathing on Aβ in the brain, which can potentially help prevent or delay the progression of AD and may contribute to the understanding of AD pathology, in particular linking Aβ production and the cholinergic system (Majdi et al., 2020).

## Data Availability

All data produced in the present study are available upon publication.

## Acknowledgments

This study was supported by Epstein Breakthrough Alzheimer’s Research Fund (PI: Mather, co-PI: Choupan) and by R01AG080652 (PI: Mather).

## Contributions

M.M. and K.N. conceptualized and designed the study with the input from R.B.C. K.N. directed the research team and analyzed the data. Data collection was performed by P.C, H.J.L, K.N and S.S. J.M., H.Y. and C.C. helped with the initial setup of the study. J.M. and H.Y. provided technical assistance with data acquisition and analyses. With the input from M.M. and K.N., N.M built a customized app and contributed to data management. L.S. performed blood assays under the supervision of E.H. K.N drafted the initial manuscript. M.M. and all other authors reviewed and edited the draft and approved the final manuscript.

