## Supplemental Information for "A randomized clinical trial reveals effects of mindfulness and slow breathing on plasma amyloid beta levels"

**Pilot Studies**

We used the following instructions shown in Supplementary Tables 1-37-9 for pilot studies 1-3.

The yellow highlights indicate changes that we made.

Supplementary Table 1.

*Meditation Instructions in Pilot Study 1*

| Belly-Focus Mindfulness Meditation | Nostril-Focus Mindfulness Meditation |
|---|---|
| **At the beginning of the session:** Welcome, please sit in a comfortable position in a chair or on a cushion with your back and neck upright and yet comfortable and gently close your eyes. If you wear glasses, feel free to take them off. | **At the beginning of the session:** Welcome, please sit in a comfortable position in a chair or on a cushion with your back and neck upright and yet comfortable and gently close your eyes. If you wear glasses, feel free to take them off. |
| Keep your mouth gently closed and focus your entire attention… entire attention on your belly. | Keep your mouth gently closed and focus your entire attention… entire attention on the area at the entrance of the nostrils. |
| Remain aware, aware of every breath coming in, every breath coming out. | Remain aware, aware of every breath coming in, every breath coming out. |
| On the inhale, allow your belly to receive your breath, on the exhale allow the belly to contract | As you inhale and exhale, simply observe changing sensations in the upper lip area as the air passes by this part of the body. |
| As you keep your attention here, notice the sensations around the belly as the breath comes in and out. Whatever sensations arise, keep your attention here, noticing the arising and passing of sensation from moment to moment. | As you keep your attention here, notice sensations at the entrance to the nostril and on the upper lip as the breath comes in and out. Whatever sensations arise, keep your attention here, noticing the arising and passing of sensation from moment to moment. |

| | |
|---|---|
| As you inhale, feel your belly expanding and as you exhale feel your belly contracting. Noticing moment to moment whatever sensations are present here in the belly. | As you inhale, feel the air moving by your upper entrance to the nostrils, on the exhale feel the slightly warmer air passing by this part of the body. Noticing moment to moment whatever sensations are present here at the entrance to the nostrils. |
| Whatever pace your breath naturally settles into is fine, no need to control it in any way. | Whatever pace your breath naturally settles into is fine, no need to control it in any way. |
| As you meditate, many thoughts, feelings, images, and memories may come up. Welcome, all that arises. Letting them rise and fall, staying with the sensations of the breath expanding and contracting the belly.<br><br>**At 5 min**: If you find yourself in conversation with a thought or feeling, simply acknowledge this is happening, meet yourself kindly, and bring attention gently back to the sensations in the belly. Sensations of the air passing in and out of the body with each passing breath. | As you meditate, many thoughts, feelings, images, and memories may come up. Welcome, all that arises. Letting them rise and fall, staying with the sensations of the breath passing by the entrance to the nostrils and the upper lip.<br><br>**At 5 min**: If you find yourself in conversation with a thought or feeling, simply acknowledge this is happening, meet yourself kindly, and bring attention gently back to the sensations at the entrance to the nostrils and above the upper lip. Sensations of the air passing in and out of the body with each passing breath. |
| **At 8 minutes:**<br>Whatever sensations arise, just observing. Observe how long the sensation lasts, how it changes with each passing moment. It may become stronger for some time, but sooner or later it passes away. No sensation is eternal, each arising and passing away. It is the same with any sensation, whether it be a tingling, an itching, warmth, coolness, or vibration, heaviness; simply observe whatever sensation is present, noticing how they change moment to moment. | **At 8 minutes:**<br>Whatever sensations arise, just observing. Observe how long the sensation lasts, how it changes with each passing moment. It may become stronger for some time, but sooner or later it passes away. No sensation is eternal, each arising and passing away. It is the same with any sensation, whether it be a tingling, an itching, warmth, coolness, or vibration, heaviness; simply observe whatever sensation is present, noticing how they change moment to moment. |

| | |
|---|---|
| **At 12 minutes:** It's quite normal for the mind to wander, if and when you notice your mind wandering, gently bring your attention back to the sensations in the belly right here and now, what sensations are present? | **At 12 minutes:** It is normal for the mind to wander, if and when you notice your mind wandering, gently bring your attention back to the sensations at the entrance to the nostrils and upper lip right here and now, what sensations are present? |
| **At 15 minutes:** Noticing where the mind is now, and gently bringing attention back to the sensations of breath and the belly. Moment to moment what sensations are present in the belly. Each moment of the inhalation … exhalation, what sensations are present now? | **At 15 minutes:** Noticing where the mind is now, and gently bringing attention back to the sensations of breath at the entrance of the nostrils. Moment to moment what sensations are present around the nostrils and upper lip. Each moment of the inhalation … exhalation, what sensations are present now? |
| **At 20 minutes:** This is the end of the session. Close your practice by setting an intention to stay connected to your body as you move slowly out of the meditation. | **At 20 minutes:** This is the end of the session. Close your practice by setting an intention to stay connected to your body as you move slowly out of the meditation. |

Supplementary Table 2.

*Meditation Instructions in Pilot Study 2*

| Belly-Focus Mindfulness Meditation | Nostril-Focus Mindfulness Meditation |
|---|---|
| As you keep your attention here, notice the sensations around the belly as the air moves in and out. As your breath flows in, feel it move down into your belly. Please observe how breathing makes your belly move. | As you keep your attention here, notice sensations at the entrance to the nostril and on the upper lip. You may feel prickles at a hair follicle or a hair moving. Please observe the sensations in this area. |

*Note.* Highlighted text was added to the instructions used in pilot study 1.

Supplementary Table 3.

*Meditation Instructions in Pilot Study 3*

| Belly-Focus Mindfulness Meditation “with” slow breathing instructions | Belly-Focus Mindfulness Meditation “without” slow breathing instructions |
|---|---|
| Take long slow breaths as I guide you. Inhale for a count to 5 and exhale for a count to 5. Breathe slowly and gently at this pace during the entire meditation exercise.<br><br>I’ll count with you for the first few breaths. Then you can count on your own. Inhale, 2, 3, 4, 5, exhale 2, 3, 4, 5. Inhale, 2, 3, 4, 5, exhale 2, 3, 4, 5. Continue breathing this way on your own.<br><br>As you keep your attention here, notice the sensations around the belly as the air moves in and out. As your breath flows in, feel it move down into your belly. Please observe how breathing makes your belly move. | As you keep your attention here, notice the sensations around the belly as the air moves in and out. As your breath flows in, feel it move down into your belly. Please observe how breathing makes your belly move. |

*Note.* Highlighted text was added to the instructions used in pilot study 2.

**Guided Meditation Scripts Used in the Actual Study**

Participants in the two meditation conditions were given the same guided meditation instructions except for the parts highlighted.

Supplementary Table 4.

*Guided Meditation Scripts*

| Mindfulness meditation with slow breathing | Mindfulness meditation without slow breathing |
|---|---|

| | |
|---|---|
| At the beginning of the session (0-4 min): Welcome, please sit in a comfortable position in a chair or on a cushion with your back and neck upright and yet comfortable and gently close your eyes. If you wear glasses, feel free to take them off. | At the beginning of the session (0-4 min): Welcome, please sit in a comfortable position in a chair or on a cushion with your back and neck upright and yet comfortable and gently close your eyes. If you wear glasses, feel free to take them off. |
| Keep your mouth gently closed and focus your entire attention… entire attention on your belly. | Keep your mouth gently closed and focus your entire attention… entire attention on your belly. |
| Remain aware, aware of every breath coming in, every breath coming out. | Remain aware, aware of every breath coming in, every breath coming out. |
| On the inhale, allow your belly to receive your breath, on the exhale allow the belly to contract. | On the inhale, allow your belly to receive your breath, on the exhale allow the belly to contract. |
| Take long slow breaths as I guide you. Inhale for a count to 5 and exhale for a count to 5. Breathe slowly and gently at this pace during the entire meditation exercise.<br><br>I'll count with you for the first few breaths. Then you can count on your own. Inhale, 2, 3, 4, 5, exhale 2, 3, 4, 5. Inhale, 2, 3, 4, 5, exhale 2, 3, 4, 5. Continue breathing this way on your own.<br><br>[pause for a few seconds before speaking again] | |

| | |
|---|---|
| Notice the sensations around the belly as the air moves in and out. As your breath flows in, feel it move down into your belly. Please observe how breathing makes your belly move. Whatever sensations arise, keep your attention here, noticing the arising and passing of sensation from moment to moment. | Notice the sensations around the belly as the air moves in and out. As your breath flows in, feel it move down into your belly. Please observe how breathing makes your belly move. Whatever sensations arise, keep your attention here, noticing the arising and passing of sensation from moment to moment. |
| As you inhale, feel your belly expanding and as you exhale feel your belly contracting. Noticing moment to moment whatever sensations are present here in the belly. | As you inhale, feel your belly expanding and as you exhale feel your belly contracting. Noticing moment to moment whatever sensations are present here in the belly. |
| As you meditate, many thoughts, feelings, images, and memories may come up. Welcome, all that arises. Letting them rise and fall, staying with the sensations of the breath expanding and contracting the belly. | As you meditate, many thoughts, feelings, images, and memories may come up. Welcome, all that arises. Letting them rise and fall, staying with the sensations of the breath expanding and contracting the belly. |
| At 5 min:<br>If you find yourself in conversation with a thought or feeling, simply acknowledge this is happening, meet yourself kindly, and bring attention gently back to the sensations in the belly. Remember to breathe slowly and gently for a count of 5 while inhaling and a count of 5 while exhaling. | At 5 min:<br>If you find yourself in conversation with a thought or feeling, simply acknowledge this is happening, meet yourself kindly, and bring attention gently back to the sensations in the belly. |

| | |
|---|---|
| At 8 minutes:<br>Whatever sensations arise, just observing. Observe how long the sensation lasts, how it changes with each passing moment. It may become stronger for some time, but sooner or later it passes away. No sensation is eternal, each arising and passing away. It is the same with any sensation, whether it be a tingling, an itching, warmth, coolness, or vibration, heaviness; simply observe whatever sensation is present, noticing how they change moment to moment. Remember to breathe slowly and gently for a count of 5 for each inhalation and a count of 5 for each exhalation during the remaining session. | At 8 minutes:<br>Whatever sensations arise, just observing. Observe how long the sensation lasts, how it changes with each passing moment. It may become stronger for some time, but sooner or later it passes away. No sensation is eternal, each arising and passing away. It is the same with any sensation, whether it be a tingling, an itching, warmth, coolness, or vibration, heaviness; simply observe whatever sensation is present, noticing how they change moment to moment. |
| At 12 minutes:<br>It's quite normal for the mind to wander, if and when you notice your mind wandering, gently bring your attention back to the sensations in the belly right here and now, what sensations are present? Remember to breathe slowly and gently. | At 12 minutes:<br>It's quite normal for the mind to wander, if and when you notice your mind wandering, gently bring your attention back to the sensations in the belly right here and now, what sensations are present? |
| At 15 minutes:<br>Noticing where the mind is now, and gently bringing attention back to the sensations of breath and the belly. Moment to moment what sensations are present in the belly. Each moment of the inhalation … exhalation, what sensations are present now? | At 15 minutes:<br>Noticing where the mind is now, and gently bringing attention back to the sensations of breath and the belly. Moment to moment what sensations are present in the belly. Each moment of the inhalation … exhalation, what sensations are present now? |
| At the end:<br>This is the end of the session. Close your practice by setting an intention to stay connected to your body as you move slowly out of the meditation. | At the end:<br>This is the end of the session. Close your practice by setting an intention to stay connected to your body as you move slowly out of the meditation. |

*Note.* Highlighted text indicates the difference between the two conditions.

**Intervention Effects on Heart Rate Variability during Practice**

We calculated log LF power, log HF power, RMSSD and heart rate averaged across all practice sessions (Supplementary Table 5). There was a significant difference between conditions during practice sessions in log LF power but not in any of the other HRV measures.

Supplementary Table 5.

*Heart Rate Variability during Practice*

| HR/HRV index | Meditation+SB *Mean (SE)* | Meditation-SB *Mean (SE)* | No Intervention *Mean (SE)* | Condition *p* |
|---|---|---|---|---|
| Log LF power (0.04-0.15 Hz) | 7.87(0.12) | 6.78(0.13) | 6.52(0.12) | <.001 |
| Log HF power (0.15-0.4 Hz) | 6.48(0.14) | 6.71(0.16) | 6.43(0.15) | 0.391 |
| RMSSD | 56.58(3.15) | 51.53(3.43) | 48.79(3.25) | 0.223 |
| Heart Rate | 79.53(1.52) | 79.58(1.65) | 78.86(1.57) | 0.936 |

**Intervention Effects on Heart rate Variability at Rest**

We examined LF power, HF power, RMSSD and mean heart rate at pre-intervention and post-intervention rest (Supplementary Table 6). There were no significant time x condition interactions for HF power, RMSSD and mean heart rate. We found a significant interaction of time and condition for LF power, $F(2,83) = 7.77$, $p < 0.001$, $\eta_p^2 = 0.158$. The interaction was driven by a significant pre-to-post increase in the M+SB condition, $t(29) = 2.215$, $p = 0.035$, and significant pre-to-post decreases in the M-SB condition, $t(25) = -2.923$, $p = 0.007$, and in the no-intervention condition, $t(29) = -2.279$, $p = 0.030$. It is possible that M+SB participants' habit of breathing slowly during daily practice carried over to the resting-state, increasing resting LF power. The subsequent analyses showing no condition differences when breathing was controlled supports this interpretation that resting-state differences between conditions in heart rate oscillations was due to differences in breathing.

Supplementary Table 6.

*Heart rate Variability at Rest*

| HR/HRV index | Meditation+SB | | Meditation-SB | | No Intervention | | Time | Time x Condition |
|---|---|---|---|---|---|---|---|---|
| | *Pre Mean (SE)* | *Post Mean (SE)* | *Pre Mean (SE)* | *Post Mean (SE)* | *Pre Mean (SE)* | *Post Mean (SE)* | *p* | *p* |
| Log LF power | 6.60 (0.19) | 7.12 (0.20) | 6.59 (0.20) | 6.19 (0.22) | 6.71 (0.19) | 6.26 (0.20) | 0.345 | <.001 |
| Log HF power | 6.84 (0.18) | 6.60 (0.18) | 6.54 (0.19) | 6.51 (0.19) | 6.68 (0.18) | 6.36 (0.18) | 0.030 | 0.393 |
| RMSSD | 56.85 (5.70) | 53.99 (3.64) | 47.64 (6.12) | 45.09 (3.91) | 50.26 (5.70) | 46.55 (3.64) | 0.247 | 0.983 |
| Heart Rate | 75.41 (1.99) | 78.09 (1.96) | 76.81 (2.14) | 79.72 (2.10) | 74.71 (1.99) | 80.49 (1.96) | <.001 | 0.402 |

**Intervention Effects on Heart Rate Variability during Controlled Breathing**

We examined LF power, HF power, RMSSD and mean heart rate while participants performed paced breathing at 15 breaths per minute at pre- and post-intervention (Supplementary Table 7). There were no significant time x condition interactions for any of the variables.

Supplementary Table 7.

*Heart Rate Variability during Controlled Breathing*

| HR/HRV index | Meditation+SB | | Meditation-SB | | No Intervention | | Time | Time x Condition |
|---|---|---|---|---|---|---|---|---|
| | *Pre Mean (SE)* | *Post Mean (SE)* | *Pre Mean (SE)* | *Post Mean (SE)* | *Pre Mean (SE)* | *Post Mean (SE)* | *p* | *p* |
| Log LF power | 6.21 (0.18) | 6.08 (0.20) | 6.11 (0.20) | 6.11 (0.23) | 5.95 (0.19) | 5.95 (0.21) | 0.614 | 0.755 |
| Log HF power | 6.75 (0.17) | 6.53 (0.16) | 6.94 (0.19) | 6.70 (0.18) | 6.62 (0.18) | 6.39 (0.16) | 0.005 | 0.996 |
| RMSSD | 55.81 (4.63) | 46.22 (3.37) | 51.07 (5.14) | 44.08 (3.74) | 44.96 (4.79) | 43.63 (3.48) | 0.011 | 0.312 |
| Heart Rate | 77.93 (2.04) | 80.86 (1.95) | 80.70 (2.27) | 83.24 (2.17) | 78.12 (2.11) | 83.14 (2.02) | 0.001 | 0.577 |

**Intervention Effects on Emotional Well-being**

The means of all emotion measures for all conditions are reported in Supplementary Table 8. In addition to the univariate analyses reported in the main text, we performed time x condition ANOVAs for each emotional well-being measure. For mood measured by POMS, there was no significant main effect of time, $F(1, 81) = 0.004$, $p = .952$, $\eta_p^2 < 0.001$, or time-by-condition interaction, $F(2, 81) = 1.24$, $p = 0.296$, $\eta_p^2 = 0.030$. For mindfulness measured by the total FFMQ score, we also did not find significant main effect of time, $F(1, 84) = 1.90$, $p = .172$, $\eta_p^2 = 0.022$, or time-by-condition interaction, $F(2, 84) = 1.68$, $p = 0.192$, $\eta_p^2 = 0.039$. Similarly for anxiety, there was no significant main effect of time, $F(1, 85) = 2.53$, $p = 0.115$, $\eta_p^2 = 0.029$, or time-by-condition interaction, $F(2, 85) = 1.39$, $p = 0.255$, $\eta_p^2 = 0.032$. For stress, there was a significant main effect of time, $F(1, 85) = 7.87$, $p = 0.006$, $\eta_p^2 = 0.085$, but no significant time-by-condition interaction, $F(2, 85) = 0.59$, $p = 0.558$, $\eta_p^2 = 0.014$. The main effect of time indicated a significant pre-to-post decrease in stress across conditions. For depression, there was a significant main effect of time, $F(1, 83) = 12.33$, $p < 0.001$, $\eta_p^2 = 0.129$, and a time-by-condition interaction, $F(2, 83) = 5.16$, $p = 0.008$, $\eta_p^2 = 0.111$. The interaction was driven by a significant pre-to-post decrease in depressive symptoms in the no-intervention condition, $t(26) = 4.324$, $p < 0.001$, but not in the meditation+SB, $t(31) = 1.871$, $p = 0.071$, or in the meditation-SB, $t(26) = -0.187$, $p = 0.853$.

Supplementary Table 8.

*Mean Emotional Well-being Score for Each Condition at Pre- and Post-intervention*

| Emotion Type | Meditation+SB | | Meditation-SB | | No Intervention | | Condition Difference at Post After Covarying Out Pre |
|---|---|---|---|---|---|---|---|
| | *Pre Mean (SE)* | *Post Mean (SE)* | *Pre Mean (SE)* | *Post Mean (SE)* | *Pre Mean (SE)* | *Post Mean (SE)* | *p* |
| Mood (POMS) | 87.39(2.14) | 84.68(2.73) | 87.15(2.29) | 89.89(2.93) | 94.73(2.34) | 94.96(2.99) | 0.203 |
| Depression (DASS) | 5.69(1.18) | 4.25(1.03) | 5.04(1.28) | 5.19(1.12) | 10.22(1.28) | 6.67(1.12) | 0.143 |
| Stress (DASS) | 10.13(1.50) | 8.44(1.37) | 10.00(1.63) | 8.74(1.49) | 14.48(1.57) | 11.38(1.44) | 0.965 |
| Anxiety (DASS) | 6.31(1.20) | 5.63(1.04) | 6.82(1.30) | 6.96(1.13) | 8.69(1.26) | 6.48(1.09) | 0.439 |
| Mindfulness (FFMQ: Total) | 41.84(1.08) | 43.44(1.08) | 42.78(1.17) | 42.44(1.18) | 39.04(1.15) | 39.57(1.16) | 0.149 |
| FFMQ Subscale (Describe) | 11.13(0.40) | 11.06(0.41) | 10.56(0.43) | 10.44(0.45) | 9.62(0.42) | 9.90(0.43) | 0.706 |
| FFMQ Subscale (Awareness) | 10.00(0.39) | 10.38(0.38) | 10.78(0.42) | 10.33(0.41) | 9.41(0.41) | 9.21(0.39) | 0.122 |
| FFMQ Subscale (Non-judging) | 11.59(0.47) | 12.19(0.40) | 11.63(0.51) | 11.96(0.44) | 10.62(0.50) | 10.69(0.42) | 0.094 |
| FFMQ Subscale (Non-reacting) | 9.13(0.41) | 9.81(0.43) | 9.82(0.45) | 9.70(0.47) | 8.52(0.43) | 9.17(0.45) | 0.587 |

**Intervention Effects on Emotional Memory**

We conducted a 3 (condition: M+SB, M-SB, no intervention) x 3 (valence of pictures: neutral, negative, positive) ANOVA for recall performance. There was a significant main effect of valence, $F(2, 162) = 18.25$, $p < 0.001$, $\eta_p^2 = 0.184$. Participants across conditions recalled more negative than neutral pictures, $t(83) = 5.850$, $p < 0.001$, more negative than positive pictures, $t(83) = 3.481$, $p < 0.001$, and more positive than neutral pictures, $t(83) = 2.604$, $p = 0.011$. For recognition memory performance, we performed 3 condition x 3 valence ANOVAs for hit rates, false alarm rates, and corrected recognition (hits minus false alarms), separately. For hits, there was a significant main effect of valence, $F(2, 168) = 18.35$, $p < 0.001$, $\eta_p^2 = 0.179$. For false alarms, there was also a significant main effect of valence, $F(2, 168) = 3.93$, $p = 0.021$, $\eta_p^2$

= 0.045. For corrected recognition, we also found a significant main effect of valence, $F(2, 162) = 8.81$, $p < 0.001$, $\eta_p^2 = 0.095$. There were no other significant findings.

**Correlation between HRV during Practice and Pre-to-post Changes in Plasma Biomarkers**

There were no significant relationships between HRV during practice and pre-to-post changes in plasma biomarkers (FDR–corrected $p < 0.05$).

Supplementary Table 9.

*Correlation between HRV during Practice and Changes in Plasma Biomarkers*

| Correlations | | Log LF-power | Log HF-power | RMSSD |
|---|---|---|---|---|
| Post-Pre Aβ40 | *r* | -0.13 | -0.13 | -0.15 |
| | *p* | 0.259 | 0.277 | 0.192 |
| | *df* | 78 | 78 | 78 |
| Post-Pre Aβ42 | *r* | -0.12 | -0.09 | -0.17 |
| | *p* | 0.313 | 0.424 | 0.146 |
| | *df* | 78 | 78 | 78 |
| Post-Pre Aβ42/40 | *r* | -0.14 | -0.20 | -0.24 |
| | *p* | 0.267 | 0.111 | 0.049 |
| | *df* | 68 | 68 | 68 |
| Post-Pre Aβ40 & Aβ42 Aggregate Z-score | *r* | -0.13 | -0.11 | -0.16 |
| | *p* | 0.255 | 0.354 | 0.160 |
| | *df* | 78 | 78 | 78 |
| Post-Pre pTau181 | *r* | 0.10 | 0.03 | 0.04 |
| | *p* | 0.382 | 0.763 | 0.693 |
| | *df* | 83 | 83 | 83 |
| Post-Pre tTau | *r* | 0.02 | 0.04 | -0.05 |
| | *p* | 0.887 | 0.756 | 0.700 |
| | *df* | 74 | 74 | 74 |
| Post-Pre pTau/tTau | *r* | 0.12 | -0.21 | -0.05 |
| | *p* | 0.317 | 0.081 | 0.673 |
| | *df* | 71 | 71 | 71 |

**Correlation between Pre-to-post Changes in HRV and Pre-to-post Changes in Plasma Biomarkers**

There were no significant correlations between pre-to-post changes in HRV and pre-to-post changes in plasma biomarkers (FDR–corrected $p < 0.05$).

Supplementary Table 10.

*Correlation between Changes in HRV and Changes in Plasma Biomarkers*

| **Correlations** | | **Post-Pre Log LF-power** | **Post-Pre Log HF-power** | **Post-Pre RMSSD** |
|---|---|---|---|---|
| Post-Pre Aβ40 | *r* | -0.11 | 0.04 | -0.05 |
| | *p* | 0.355 | 0.705 | 0.667 |
| | *df* | 75 | 75 | 75 |
| Post-Pre Aβ42 | *r* | -0.13 | 0.04 | -0.09 |
| | *p* | 0.273 | 0.757 | 0.447 |
| | *df* | 75 | 75 | 75 |
| Post-Pre Aβ42/40 | *r* | -0.12 | -0.12 | -0.09 |
| | *p* | 0.330 | 0.351 | 0.459 |
| | *df* | 65 | 65 | 65 |
| Post-Pre Aβ40 & Aβ42 Aggregate Z-score | *r* | -0.14 | 0.03 | -0.09 |
| | *p* | 0.239 | 0.775 | 0.425 |
| | *df* | 75 | 75 | 75 |
| Post-Pre pTau181 | *r* | -0.04 | 0.00 | -0.02 |
| | *p* | 0.759 | 0.992 | 0.836 |
| | *df* | 80 | 80 | 80 |
| Post-Pre tTau | *r* | -0.01 | -0.14 | -0.16 |
| | *p* | 0.925 | 0.237 | 0.193 |
| | *df* | 71 | 71 | 71 |
| Post-Pre pTau/tTau | *r* | 0.02 | 0.16 | 0.18 |
| | *p* | 0.887 | 0.197 | 0.140 |
| | *df* | 68 | 68 | 68 |

**Supplementary Fig. 1.**

*Heart Rate Power Spectral Density in Pilot Study 1 (a), Pilot Study 2 (b) and Pilot Study 3 (c)*

**a) Pilot study 1**

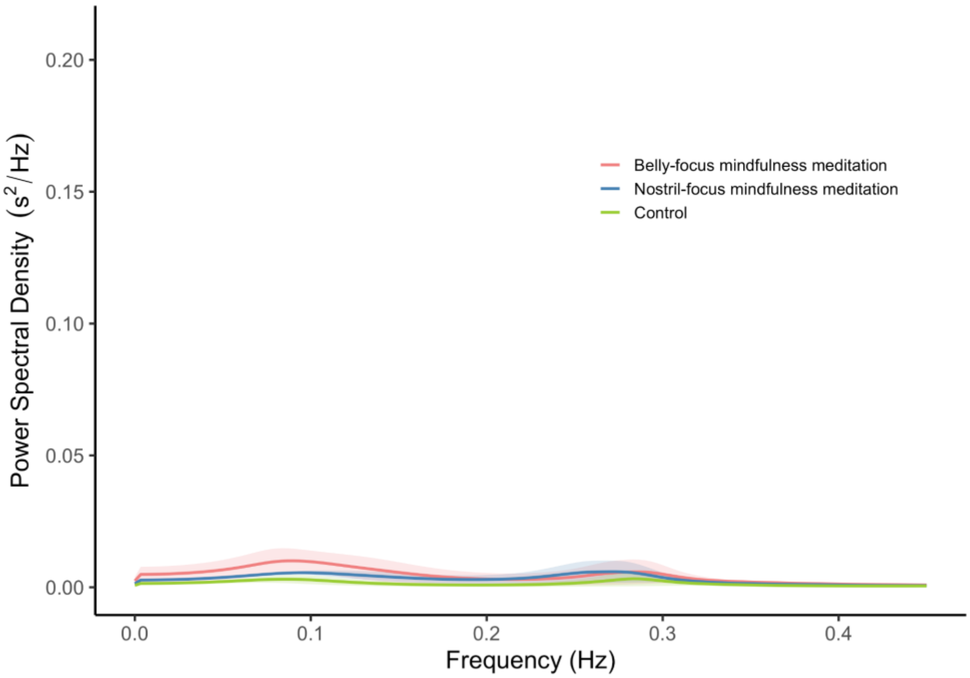


**b) Pilot study 2**

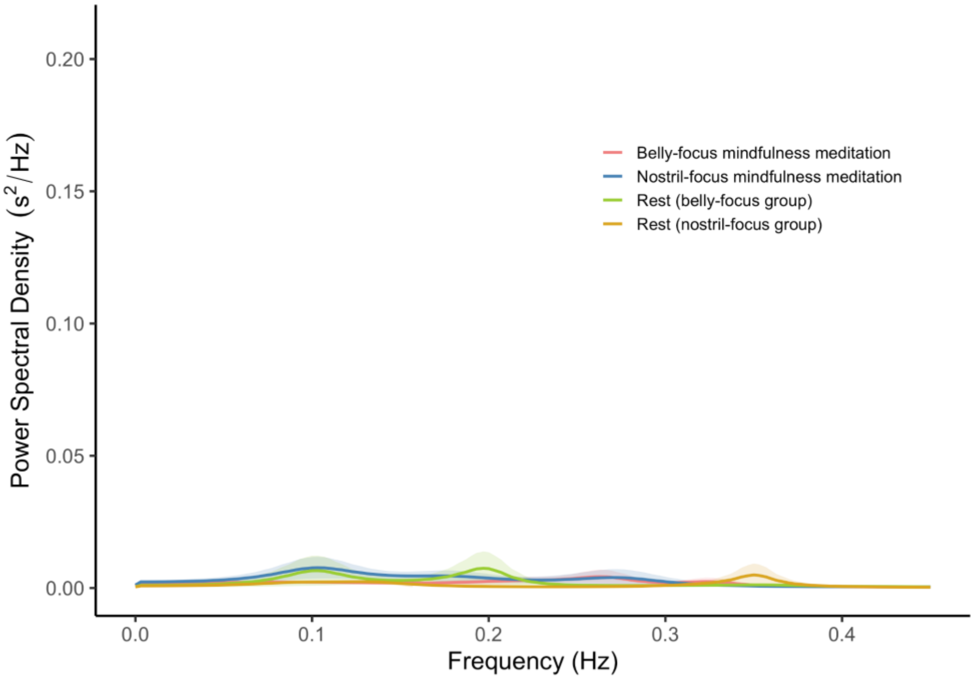


**c) Pilot study 3**

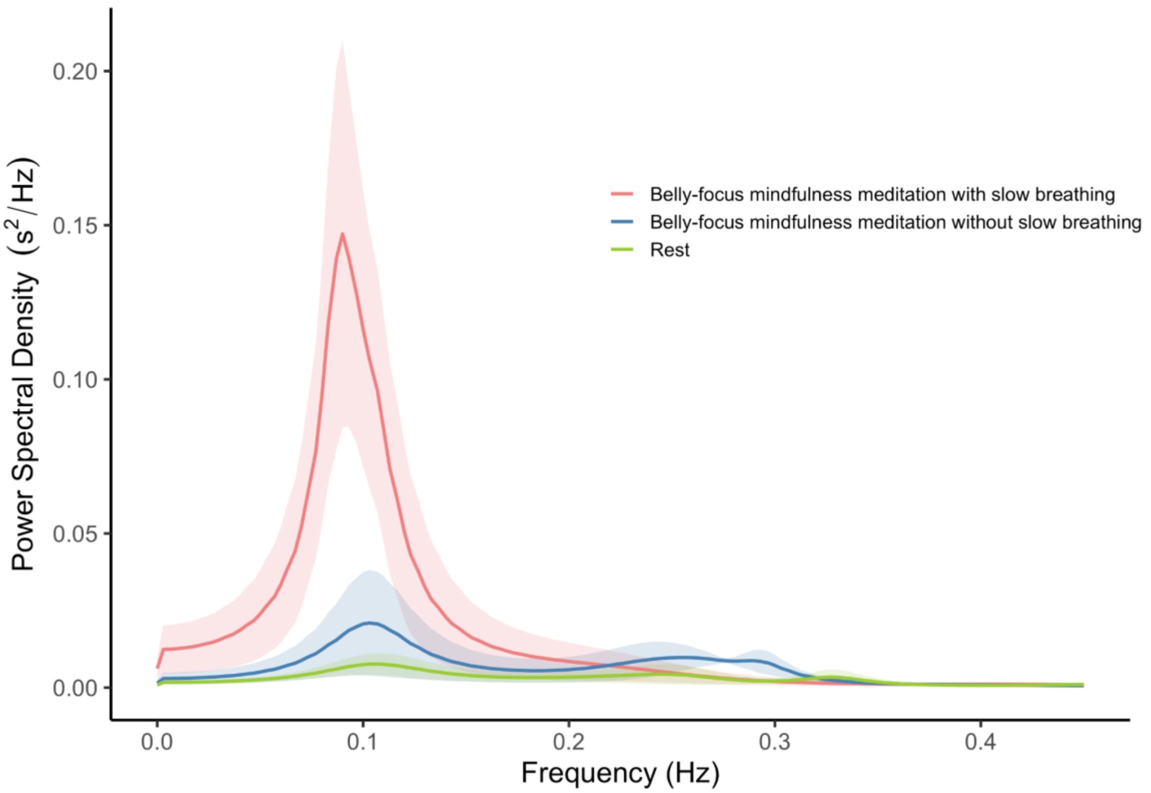


*Note.* Neither belly-focus or nostril-focus meditation condition increased LF-HRV relative to control (a) or rest conditions (b). The belly-focus meditation group who received explicit slow breathing instructions increased LF-HRV relative to rest, which was not observed in the belly-focus meditation group without slow breathing instructions (c).
